# Genome-wide association studies reveal novel locus with sex-/therapy-specific fracture risk effects in childhood cancer survivors

**DOI:** 10.1101/2020.09.21.20196121

**Authors:** Cindy Im, Nan Li, Wonjong Moon, Qi Liu, Lindsay M. Morton, Wendy M. Leisenring, Rebecca M. Howell, Eric J. Chow, Charles A. Sklar, Carmen L. Wilson, Zhaoming Wang, Yadav Sapkota, Wassim Chemaitilly, Kirsten K. Ness, Melissa M. Hudson, Leslie L. Robison, Smita Bhatia, Gregory T. Armstrong, Yutaka Yasui

## Abstract

Survivors of childhood cancer treated with radiation therapy (RT) and osteotoxic chemotherapies are at increased risk for fractures. However, research focusing on how genetic and clinical susceptibility factors jointly contribute to fracture risk among long-term (≥5 years) survivors of childhood cancer has been limited. To address this gap, we conducted genome-wide association studies of fracture risk in 2,453 participants from the Childhood Cancer Survivor Study (CCSS) using Cox regression models and prioritized sex- and treatment-stratified genetic associations. Replication analyses were conducted in an independent survivor sample from the St. Jude Lifetime Cohort Study (SJLIFE). We identified a genome-wide significant (P<5⨯10^−8^) fracture risk locus, 16p13.3 (*HAGHL*), among female CCSS survivors (N=1,289) with strong evidence of sex-specific effects (P_sex-heterogeneity_<7⨯10^−6^). We found rs1406815 showed the strongest association with fracture risk after replication (HR_meta-analysis_ per risk allele=1.43, P=8.2⨯10^−9^; N=1,935 women). While the association between rs1406815 and fracture risk was weak among female survivors who did not receive radiation therapy (RT) (HR_CCSS_=1.22, P=0.11), the association strength increased with greater RT doses to the head or neck (HR_CCSS_=1.88, P=2.4⨯10^−10^ in those with any head/neck RT; HR_CCSS_=3.79, P=9.1⨯10^−7^ in those treated with >36 Gray). *In silico* bioinformatics analyses suggest these fracture risk alleles regulate *HAGHL* gene expression and related bone resorption pathways, and are plausibly moderated by head/neck RT. Genetic risk profiles integrating this locus may help identify young female survivors who would benefit from targeted interventions to reduce fracture risk.

## INTRODUCTION

Childhood cancer survivors are at increased risk for developing bone-related late effects. Treatment with osteotoxic chemotherapies (e.g., corticosteroids, methotrexate) may adversely affect normal bone metabolism and skeletal development, while radiation therapy (RT) can induce bone tissue damage and endocrinopathies that influence bone loss.^(1,2)^ Other factors contributing to bone fragility risk include malignancy-related pathologies (e.g., leukemia) and deficiencies in childhood physical activity and nutrition.^(1,2)^ These clinical factors are hypothesized to disrupt the acquisition of sufficient peak bone mass during childhood and adolescence, elevating risk for early onset osteoporosis and subsequent fractures in survivors.^(1,2)^

In the general population, fractures are primarily responsible for increased morbidity and mortality risk among individuals with osteoporosis.^(3,4)^ As such, reduced bone mineral density (BMD) is a significant public health concern, as BMD deficits are the best clinical predictor of future fractures. Previous fracture history^(5,6)^ and biological sex^(7,8)^ are also critical determinants of fracture risk. In the largest-to-date study of adults treated for childhood cancer (N=1,713),^(9)^ the prevalence of BMD deficits consistent with a diagnosis of osteoporosis (*Z*-score≤-2.5 SD) was estimated to be ∼10% among survivors with a median age of 32 years, similar to the prevalence of osteoporosis among adults in the United States aged 60-80 years.^(10)^ Prevalence estimates of BMD deficits in smaller studies of long-term survivors have been reported to be as high as 30% to 50%, varying by diagnostic group, time from diagnosis, and treatment exposures.^(1,2,11-14)^ While broader studies of incident fractures after childhood cancer diagnosis are limited, studies of acute lymphoblastic leukemia survivors have reported higher incident fracture rates during and immediately after treatment: one study observed fracture rates (at any skeletal site) to be ∼six-fold higher during the three-year follow-up period after diagnosis relative to healthy controls,^(15)^ while another reported a four-year vertebral fracture cumulative incidence of ∼26%.^(16)^ Recent studies of survivors have also reported differential risk for BMD deficits^(11,17)^ and fractures^(18)^ by sex and treatment exposures, which may reflect increased sex-specific vulnerabilities to certain therapeutic agents during childhood and adolescence.^(11)^

Given that the risk for BMD deficits and fractures after treatment for childhood cancer varies among survivors with similar clinical risk profiles, insight into the joint genetic and clinical determinants of fracture risk in survivors is needed. Published genome-wide association studies (GWAS) of BMD in the general population^(19,20)^ have identified hundreds of associated genetic loci, while GWAS of fracture risk^(19,21)^ have detected over a dozen. To our knowledge, however, no fracture risk GWAS have been conducted in childhood cancer survivors. Therefore, we performed GWAS of incident fracture risk after diagnosis in 2,453 participants of the Childhood Cancer Survivor Study (CCSS). To evaluate the modifying effects of sex and cancer therapies known to adversely affect bone, we conducted sex-specific GWAS and evaluated treatment-stratified genetic associations.

## MATERIALS AND METHODS

### Survivor cohorts and fracture

For discovery analyses, we evaluated data from CCSS, the largest multi-institutional cohort of long-term (≥5 years) survivors of childhood cancer in North America. Survivors were diagnosed before age 21 years between 1970 and 1986, with prospective follow-up of late effects through periodic surveys.^(22-24)^ In this study, we included CCSS participants with DNA genotype data who responded to a follow-up questionnaire requesting detailed lifetime fracture histories (Supplemental Methods). Participants with bone tumor primary diagnoses and allogeneic stem cell transplantation history were excluded. Qualifying incident first fractures after primary cancer diagnosis were identified (Supplemental Methods). We considered fractures at any skeletal site, given traumatic fractures and fractures at sites beyond the hip/spine/wrist are also predicted by low BMD.^(3)^ Covariate data were abstracted from medical records or self-reported in surveys (Supplemental Methods).

Replication analyses were performed with data from SJLIFE,^(25,26)^ a retrospectively- constructed cohort study of 5-year survivors treated for pediatric cancer at St. Jude Children’s Research Hospital (SJCRH) with prospective medical assessment of late effects. Criteria applied to exclude participants and define qualifying fractures in discovery analyses were applied in replication analyses. Most (84.3%) fracture histories were taken from medical history interviews conducted by clinicians at SJCRH visits; otherwise, self-reported responses to fracture prompts identical to the CCSS questionnaires were used. Data for other covariates were clinically assessed during SJCRH visits or abstracted from medical records (Supplemental Methods).

All CCSS and SJLIFE study protocols and contact documents were approved by the institutional review boards of participating study institutions. All study participants provided informed consent. A flow diagram summarizing inclusion criteria for discovery and replication study participation is provided in Supplemental Figure 1.

### Genotype data

Methods used to generate genotype data in CCSS and SJLIFE have been described elsewhere.^(27-30)^ In brief, DNA was genotyped using the Illumina HumanOmni5Exome array and imputed using Minimac3^(31)^ for CCSS samples, while whole genome sequencing was performed using the Illumina HiSeq X10 platform with an average coverage per sample of 36.8X in SJLIFE. Stringent sample and variant quality control was applied and analyses were restricted to participants of European genetic ancestry in CCSS and SJLIFE (Supplemental Methods). Discovery analyses in CCSS were performed with ∼5.4 million SNPs with minor allele frequency ≥5% and high imputation quality scores (r^2^≥0.8).

### Statistical analysis Discovery in CCSS

To estimate the additive effects of each SNP allele on fracture risk following diagnosis, Cox proportional hazards models with age as the time scale were used, adjusting for ancestry (first 10 principal components), sex, attained height and weight, premature menopause status, and treatments determined to be relevant through univariate association testing (Supplemental Methods): exposure to corticosteroids; intravenous (IV) methotrexate dose; intrathecal (IT) methotrexate dose; and maximum tumor dose (maxTD) from RT to any of seven major body regions (head, neck, chest, abdomen, pelvis, arm, leg).^(32)^ Associations with P<5⨯10^−8^ from two-sided tests were considered to be genome-wide significant. Descriptive cumulative incidence curves among the CCSS survey respondents were examined to compare unadjusted fracture risk by years of follow-up across SNP-genotype groups. Genomic region plots surrounding genome-wide significant SNPs were generated with LocusZoom software.^(33)^ Testing for sex-heterogeneous effects was conducted with GWAMA v2.2.2^(34)^ for suggestively significant (P<1⨯10^−5^) SNPs from discovery (Supplemental Methods).

### Replication in SJLIFE and meta-analysis

Genome-wide significant SNP-fracture associations in CCSS were evaluated for replication in SJLIFE, using the same statistical model from discovery analyses. Associations with replication P≤0.05 (two-sided) with association directions consistent with discovery were considered replicated. For replicated SNP associations, summary effect estimates combining CCSS and SJLIFE association results were computed using the fixed-effects inverse variance- weighted meta-analysis method with GWAMA v2.2.2.^(34)^ The Cochran’s Q statistic and I^2^ inconsistency index were assessed for effect heterogeneity.

### Cancer therapy modifiers of SNP effects

Replicated SNP associations were re-evaluated in treatment-stratified analyses under three composite treatment definitions: head/neck radiation (maxTD for head or neck), trunk radiation (maxTD for chest, abdomen, or pelvis), and chemotherapy (any corticosteroid exposure and IV or IT methotrexate dose). Given the high prevalence of endocrinopathies after RT to the cranial, hypothalamic-pituitary, or neck regions^(35)^, we evaluated estimates stratified by head/neck RT dose separately from trunk RT. We compared adjusted SNP associations in strata with no, any, >medium, and >high dose exposures for these treatment definitions, with medium and high dose corresponding to median and 3^rd^ quartile doses in CCSS, respectively.

### Functional/regulatory annotation of SNPs in credible sets

Adopting an annotation procedure similar to Gaulton *et al*.^(36)^, we constructed 99% credible intervals or SNP sets with 99% probability of containing the causal variant using a Bayesian approach^(36-38)^ for each replicated locus (Supplemental Methods) for annotation, given that: (a) the most strongly associated SNP may not directly influence fracture risk; (b) >1 causal variant may be present at a locus; and (c) the signal could reflect the effects of complex genetic variation, e.g., haplotypes. We interrogated credible-set SNP associations in recent GWAS of bone-related phenotypes^(19,20)^ and phenome-wide association studies^(39)^ (PheWAS), and functional/regulatory annotations using external genomic data resources (Supplemental Methods). Lastly, we tested whether credible-set SNPs were likely to drive fracture risk signals in survivors through promoter regulatory mechanisms in specific cell types by using an enrichment test procedure.^(36)^ Specifically, we compared the observed mean posterior probability of credible-set SNPs directly overlapping promoter regions^(40)^ to its null distribution generated with 100,000 randomly-shifted promoter region annotations across cell types specified *a priori* for relevance to fracture risk in survivors and comparison cell types from the Encyclopedia of DNA Elements^(41)^ (ENCODE) Project (Supplemental Methods).

## RESULTS

### Genetic variants at *HAGHL, CD86* loci are associated with fracture risk in female survivors

In the CCSS discovery cohort (N=2,453), the median age at follow-up survey completion was 42 years (IQR=36-48 years) and ≥1 post-diagnosis fracture was reported by 37.9% of survivors (930 incident fractures) (Supplemental Tables 1-2). Male survivors had significantly greater unadjusted risk of fracture after diagnosis (40.6% in males versus 29.3% in females at 30 years post-diagnosis, P=2.0⨯10^−7^; Supplemental Figure 2). Limb fractures accounted for the majority (>60%) of post-diagnosis fractures (Supplemental Table 3). While cancer treatment exposures were symmetrically distributed between sexes (Supplemental Table 4), increases in post-diagnosis fracture risk were associated with increasing IV and IT methotrexate dose in male survivors (Supplemental Table 5) and higher RT dosages in female survivors (Supplemental Figure 3).

Analyses of the ∼5.4 million common autosomal SNPs in the sex-combined and male-specific CCSS discovery samples yielded no genome-wide significant (P<5⨯10^−8^) associations with post-diagnosis fracture risk (Supplemental Figure 4). In the female-specific CCSS discovery sample (N=1,289), we identified three SNPs at the *CD86* locus (3q13.33) and four SNPs at the *HAGHL* locus (16p13.3) with fracture risk associations meeting the genome-wide significance threshold (Table 1). The genomic inflation factor across all analyses was ≤1.03, indicating adequate control of potential population stratification confounding. Global comparisons of SNP associations with post-diagnosis fracture risk by sex are provided in side-by-side Manhattan plots (Figure 1) and quantile-quantile (QQ) plots (Supplemental Figure 5). We observed no overlap between genome-wide significant variants in CCSS and previously reported fracture risk susceptibility loci, with all discovered SNPs located >1 Mb from lead SNPs in published fracture GWAS (Figure 1, Supplemental Table 6).

**Table 1:** Sex-heterogeneous SNP effects on post-diagnosis fracture risk in childhood cancer survivors from the CCSS discovery cohort

|  |  |  |  |  | Women<br>(N=1,289) |  |  | Men<br>(N=1,164) |  |  | Sex heterogeneity<sup>a</sup> |  |
| --- | --- | --- | --- | --- | --- | --- | --- | --- | --- | --- | --- | --- |
| SNP | Chr | BP | EA | NEA | EAF | HR (95% CI) | P | EAF | HR (95% CI) | P | Effects | P<sub>sex-het</sub> |
| rs4315642 | 3 | 121836049 | C | T | 0.28 | 0.64 (0.54 to 0.75) | 4.1x10<sup>-8</sup> | 0.29 | 1.08 (0.95 to 1.24) | 0.24 | -+ | 8.1x10<sup>-7</sup> |
| rs2681399 | 3 | 121835908 | G | A | 0.28 | 0.64 (0.54 to 0.75) | 4.7x10<sup>-8</sup> | 0.29 | 1.09 (0.95 to 1.25) | 0.23 | -+ | 7.9x10<sup>-7</sup> |
| rs2681400 | 3 | 121837377 | T | C | 0.28 | 0.64 (0.54 to 0.75) | 4.7x10<sup>-8</sup> | 0.29 | 1.08 (0.95 to 1.24) | 0.25 | -+ | 9.6x10<sup>-7</sup> |
| rs12448432 | 16 | 778820 | A | G | 0.19 | 1.55 (1.33 to 1.81) | 1.2x10<sup>-8</sup> | 0.20 | 0.91 (0.78 to 1.07) | 0.26 | + - | 2.2x10<sup>-6</sup> |
| rs1406815 | 16 | 778158 | G | C | 0.19 | 1.55 (1.33 to 1.80) | 1.5x10<sup>-8</sup> | 0.21 | 0.92 (0.78 to 1.07) | 0.28 | + - | 3.0x10<sup>-6</sup> |
| rs9928077 | 16 | 784765 | T | C | 0.19 | 1.54 (1.32 to 1.79) | 2.6x10<sup>-8</sup> | 0.21 | 0.92 (0.78 to 1.07) | 0.28 | + - | 3.9x10<sup>-6</sup> |
| rs12597563 | 16 | 787738 | C | G | 0.17 | 1.57 (1.34 to 1.84) | 2.8x10<sup>-8</sup> | 0.18 | 0.91 (0.77 to 1.08) | 0.29 | + - | 4.1x10<sup>-6</sup> |

a. For sex heterogeneity testing, a Bonferroni-corrected p-value threshold was used ( $P < 0.05 / [41 \text{ evaluated SNPs with suggestive significance in sex-specific discovery analyses}] = 1.2 \times 10^{-3}$ ) to assess statistical significance. The direction of the fracture risk associations by sex are provided in the “Effects” column, with results in female survivors presented first [left] followed by results in male survivors [right]; “-” corresponds to decreasing risk and “+” corresponds to increasing risk.
Abbreviations: SNP, single nucleotide polymorphism; Chr, chromosome; BP, base position, GRCh37 (hg19) build; EA, effect (risk) allele; NEA, non-effect (reference) allele; EAF, effect allele frequency; HR, hazard ratio; CI, confidence interval; P<sub>sex-het</sub>, sex heterogeneity test p-value.

**Figure 1:**
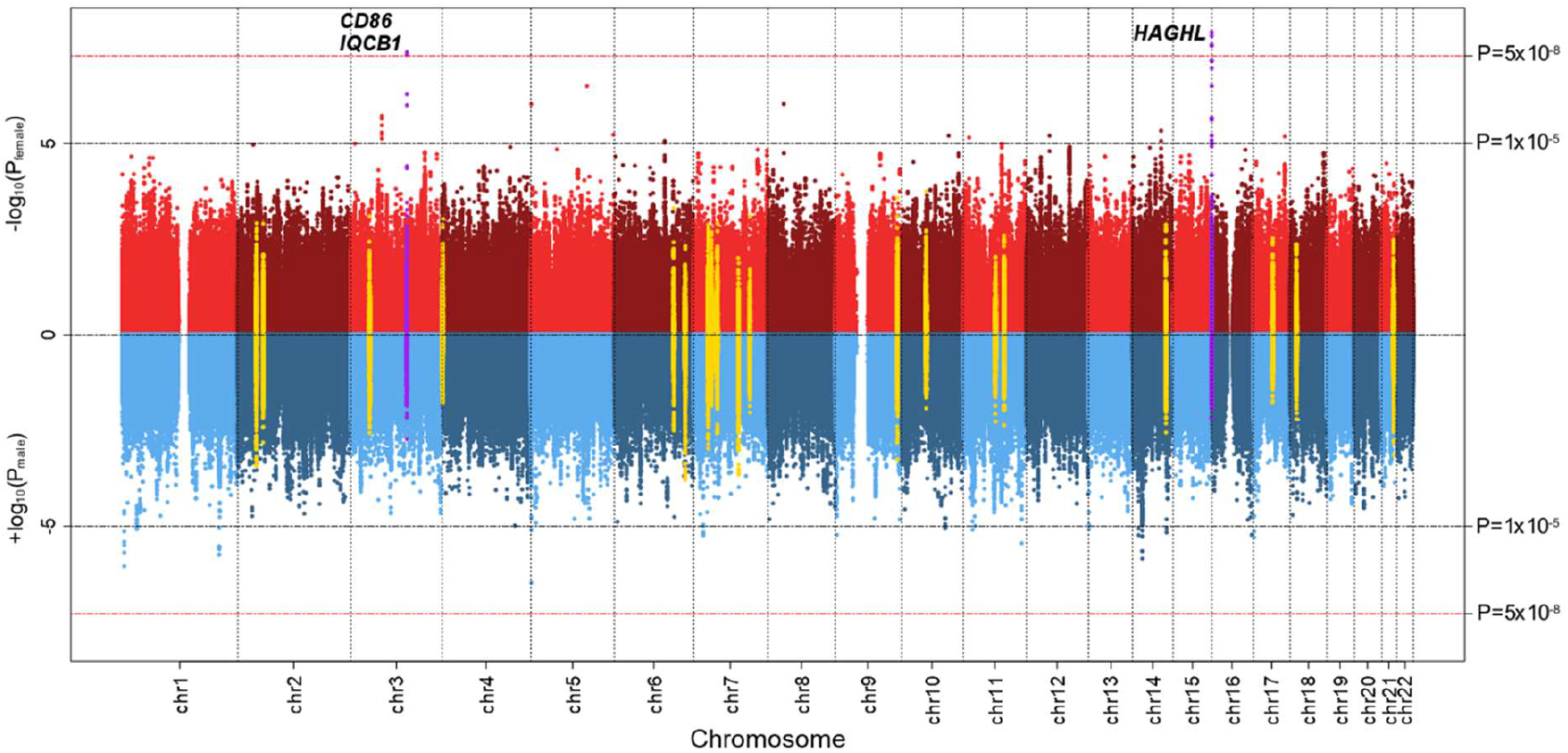
Sex-specific plots of CCSS discovery analysis p-values for autosomal SNP associations with post-diagnosis fracture risk in 2,453 childhood cancer survivors. On top (red) is the Manhattan plot of –log_10_ p-values (y-axis) by SNP genomic position (x-axis) from the genome- wide association analysis in 1,289 female survivors. On bottom (blue) is the inverted Manhattan plot (+log10 p-values) from the corresponding analysis in 1,164 male survivors. The red dashed horizontal line signifies the genome-wide significance threshold (P<5×10^−8^ from two-sided statistical tests). Log_10_ p-values for SNPs with previously reported genome-wide significant associations with fracture risk and nearby SNPs (50-kb window) are depicted in yellow. Sex- specific log_10_ p-values for genome-wide significant loci in female survivors are shown in purple.

Regional plots of SNP associations with subsequent fracture risk at the *CD86* and *HAGHL* loci in female CCSS survivors are provided in Supplemental Figures 6-7. The genome-wide significant SNPs at the *CD86* locus are in high linkage disequilibrium (LD r^2^>0.99, 1000G EUR), and rs4315642 (sample effect allele frequency or EAF=0.28) had the strongest association (HR=0.64, 95% CI: 0.54-0.75, P=4.1⨯10^−8^) (Table 1). Among the high LD (r^2^≥0.84, 1000G EUR) genome-wide significant SNPs at the *HAGHL* locus, the most significantly associated SNP was rs12448432 (EAF=0.20; HR=1.55, 95% CI: 1.33-1.81, P=1.2⨯10^−8^). All seven SNPs were characterized by significant allelic effect heterogeneity by sex (Bonferroni-corrected threshold P<0.05/[41 evaluated SNPs]=1.2⨯10^−3^; Table 1).

### Replication of female-specific *HAGHL* SNP associations with fracture risk

We evaluated all genome-wide significant SNP associations in an independent sample of survivors from SJLIFE (N=1,417; 46.0% reported post-diagnosis fractures). Among female SJLIFE survivors (N=646), we replicated three of the four *HAGHL* SNP associations with increased post-diagnosis fracture risk (HR=1.23-1.24, P≤0.05, Table 2). Meta-analysis with both female survivor cohorts (N=1,935) revealed rs1406815 (chr16:778158, GRCh37) had the strongest association with fracture risk (HR=1.43, 95% CI: 1.27-1.62, P=8.2⨯10^−9^). Inverse associations of *HAGHL* SNPs with fracture risk seen in male CCSS survivors were also observed in SJLIFE, with larger protective effects and statistical significance (N=771, P<0.01). *CD86* locus SNPs did not replicate, showing the opposite direction of association from discovery.

**Table 2:** Replication results for genome-wide significant sex-heterogeneous SNP associations with post-diagnosis fracture in survivors from the SJLIFE cohort and meta-analysis of replicated (P≤0.05) associations

|  |  |  |  |  | CCSS Discovery<br>(N=1,289 women) |  |  | SJLIFE Replication<br>(N=646 women) |  |  | Meta-analysis<br>(N=1,935 women) |  |  |  |  | SJLIFE Replication<br>(N=771 men) |  |  |
| --- | --- | --- | --- | --- | --- | --- | --- | --- | --- | --- | --- | --- | --- | --- | --- | --- | --- | --- |
| SNP | Chr | BP | EA | NEA | EAF | HR<br>(95% CI) | P | EAF | HR<br>(95% CI) | P | EAF | HR<br>(95% CI) | P | $P_{het}$ | $I^2$ | EAF | HR<br>(95% CI) | P |
| rs4315642 | 3 | 121836049 | C | T | 0.28 | 0.64<br>(0.54 to 0.75) | $4.1 \times 10^{-8}$ | 0.28 | 1.24<br>(1.01 to 1.52) | 0.04 | -- | -- | -- | -- | -- | 0.26 | 0.91<br>(0.77 to 1.07) | 0.26 |
| rs2681399 | 3 | 121835908 | G | A | 0.28 | 0.64<br>(0.54 to 0.75) | $4.7 \times 10^{-8}$ | 0.28 | 1.26<br>(1.02 to 1.55) | 0.03 | -- | -- | -- | -- | -- | 0.26 | 0.91<br>(0.78 to 1.08) | 0.28 |
| rs2681400 | 3 | 121837377 | T | C | 0.28 | 0.64<br>(0.54 to 0.75) | $4.7 \times 10^{-8}$ | 0.28 | 1.25<br>(1.02 to 1.54) | 0.03 | -- | -- | -- | -- | -- | 0.26 | 0.90<br>(0.76 to 1.06) | 0.22 |
| rs12448432 | 16 | 778820 | A | G | 0.19 | 1.55<br>(1.33 to 1.81) | $1.2 \times 10^{-8}$ | 0.21 | 1.23<br>(1.00 to 1.51) | 0.05 | 0.20 | 1.43<br>(1.27 to 1.62) | $9.1 \times 10^{-9}$ | 0.08 | 0.68 | 0.19 | 0.78<br>(0.64 to 0.93) | 0.01 |
| rs1406815 | 16 | 778158 | G | C | 0.19 | 1.55<br>(1.33 to 1.80) | $1.5 \times 10^{-8}$ | 0.21 | 1.24<br>(1.01 to 1.53) | 0.04 | 0.20 | 1.43<br>(1.27 to 1.62) | $8.2 \times 10^{-9}$ | 0.09 | 0.65 | 0.20 | 0.74<br>(0.62 to 0.90) | $1.9 \times 10^{-3}$ |
| rs9928077 | 16 | 784765 | T | C | 0.19 | 1.54<br>(1.32 to 1.79) | $2.6 \times 10^{-8}$ | 0.21 | 1.23<br>(1.00 to 1.52) | 0.05 | 0.20 | 1.43<br>(1.26 to 1.61) | $1.6 \times 10^{-8}$ | 0.09 | 0.65 | 0.20 | 0.75<br>(0.63 to 0.91) | $2.8 \times 10^{-3}$ |
| rs12597563 | 16 | 787738 | C | G | 0.17 | 1.57<br>(1.34 to 1.84) | $2.8 \times 10^{-8}$ | 0.18 | 1.17<br>(0.94 to 1.46) | 0.17 | -- | -- | -- | -- | -- | 0.18 | 0.76<br>(0.63 to 0.93) | 0.01 |

Abbreviations: SNP, single nucleotide polymorphism; Chr, chromosome; BP, base position, GRCh37 (hg19) build; EA, effect (risk) allele; NEA, non-effect (reference) allele; EAF, effect allele frequency; HR, hazard ratio; CI, confidence interval; $P_{het}$ , p-value from Cochran's Q test for effect heterogeneity; $I^2$ , inconsistency index.

### *HAGHL* SNP effects on fracture risk increase with previous head/neck radiation therapy

In treatment-stratified analyses, we found that strata with increasing doses of head/neck RT showed corresponding increases in post-diagnosis fracture risk associations with the *HAGHL* SNPs in both survivor cohorts, while strata with increasing trunk RT and composite chemotherapy doses did not (Figure 2, Supplemental Table 7). For example, among female survivors with no exposure to head/neck radiation in CCSS, the association between rs1406815 and fracture risk was weak (HR=1.22, P=0.11), increasing with any exposure (HR=1.88, P=2.4⨯10^−10^), and was appreciably greater among those with high head/neck radiation exposures (>3^rd^ quartile dose or 36 Gy stratum HR=3.79, P=9.1⨯10^−7^). Similar magnitudes of association between rs1406815 and fracture risk were seen among female survivors in SJLIFE (no head/neck radiation HR=1.38, P=0.03; >36 Gy head/neck RT HR=3.08, P=0.03). Among female CCSS survivors with any head/neck RT and homozygous rs1406815 risk alleles, the cumulative incidence of fracture was 37.1% and 60.0% at 15 years and 30 years post-diagnosis, respectively (Figure 3A). No comparable increases in fracture risk was observed among male survivors with identical genetic and treatment risk profiles (Figure 3B).

**Figure 2:**
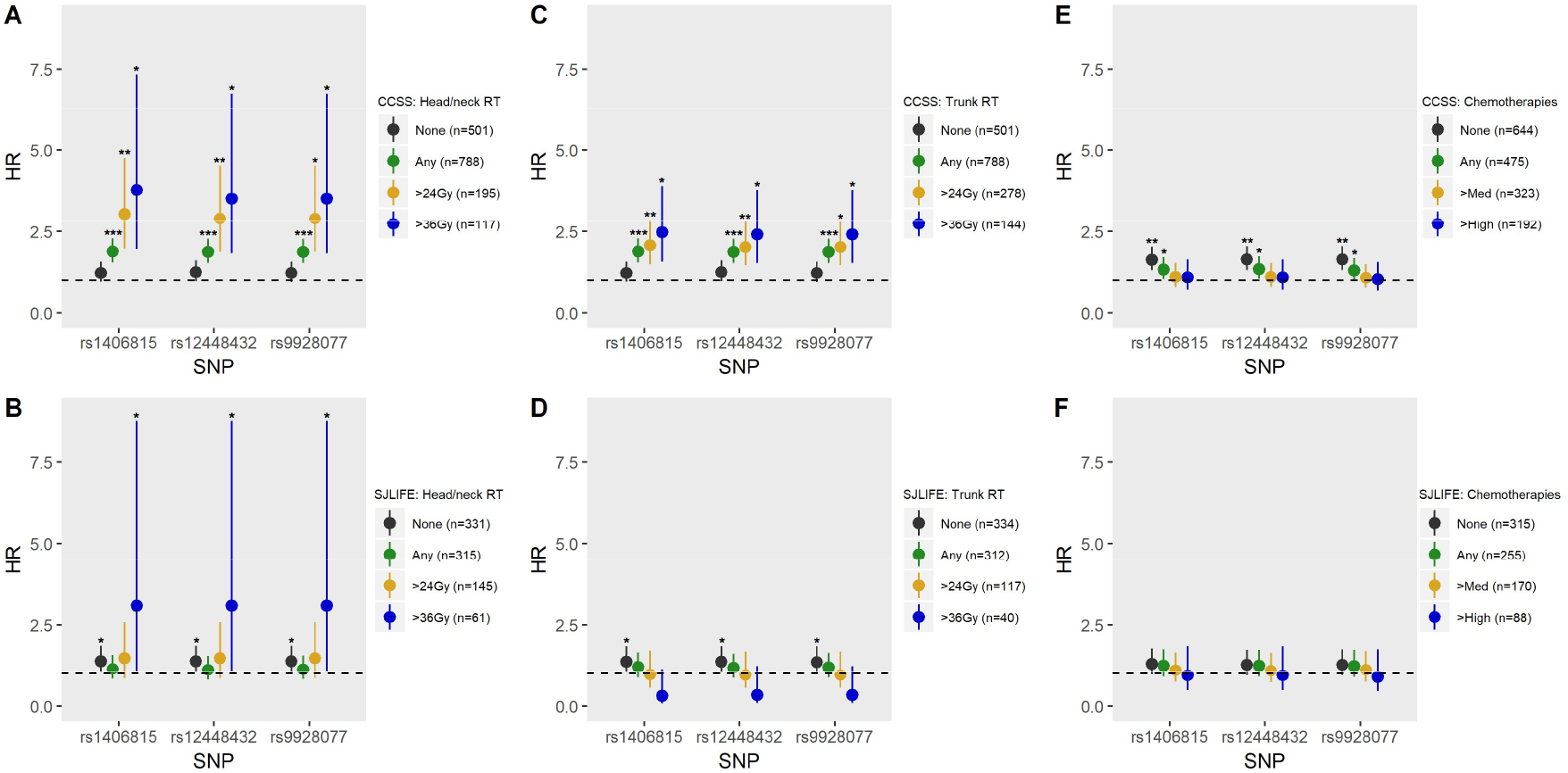
Cancer treatment-stratified associations between replicated *HAGHL* locus SNPs and post-diagnosis fracture risk in female survivors from CCSS and SJLIFE. Top row plots (2A, 2C, 2E) show stratum-specific HRs (dots) and 95% confidence intervals (whiskers) for associations between post-diagnosis fracture risk and risk alleles for each of the three replicated *HAGHL* locus SNPs (rs1406815, rs12448432, rs9928077) in CCSS female survivors (N=1,289), while bottom row plots (2B, 2D, 2F) show corresponding estimates in SJLIFE female survivors (N=646). Stratum-specific HRs for having been exposed to head/neck radiation therapy (RT), trunk RT, and chemotherapy are shown in the left (2A, 2B), middle (2C, 2D), and right columns (2E, 2F), respectively. Statistical significance thresholds for p-values from two-sided tests for genome-wide (shown as ***, for P<5×10^−8^), suggestive (shown as **, for 5⨯10^−8^≤P<5×10^−5^), and nominal (shown as *, for 5⨯10^−5^≤ P<0.05) significance are provided for stratum-specific HRs. The dashed line signifies HR=1. For 2E and 2F, the following exposures to corticosteroids and methotrexate were considered for having been exposed to the relevant chemotherapy: “any” exposure; “medium (med)” exposure as defined by any exposure to corticosteroids and greater than median IV or IT methotrexate dose; and “high” exposure as defined by any exposure to corticosteroids and greater than third quartile IV or IT methotrexate dose. The number of survivors in each treatment stratum are provided in plot legends.

**Figure 3:**
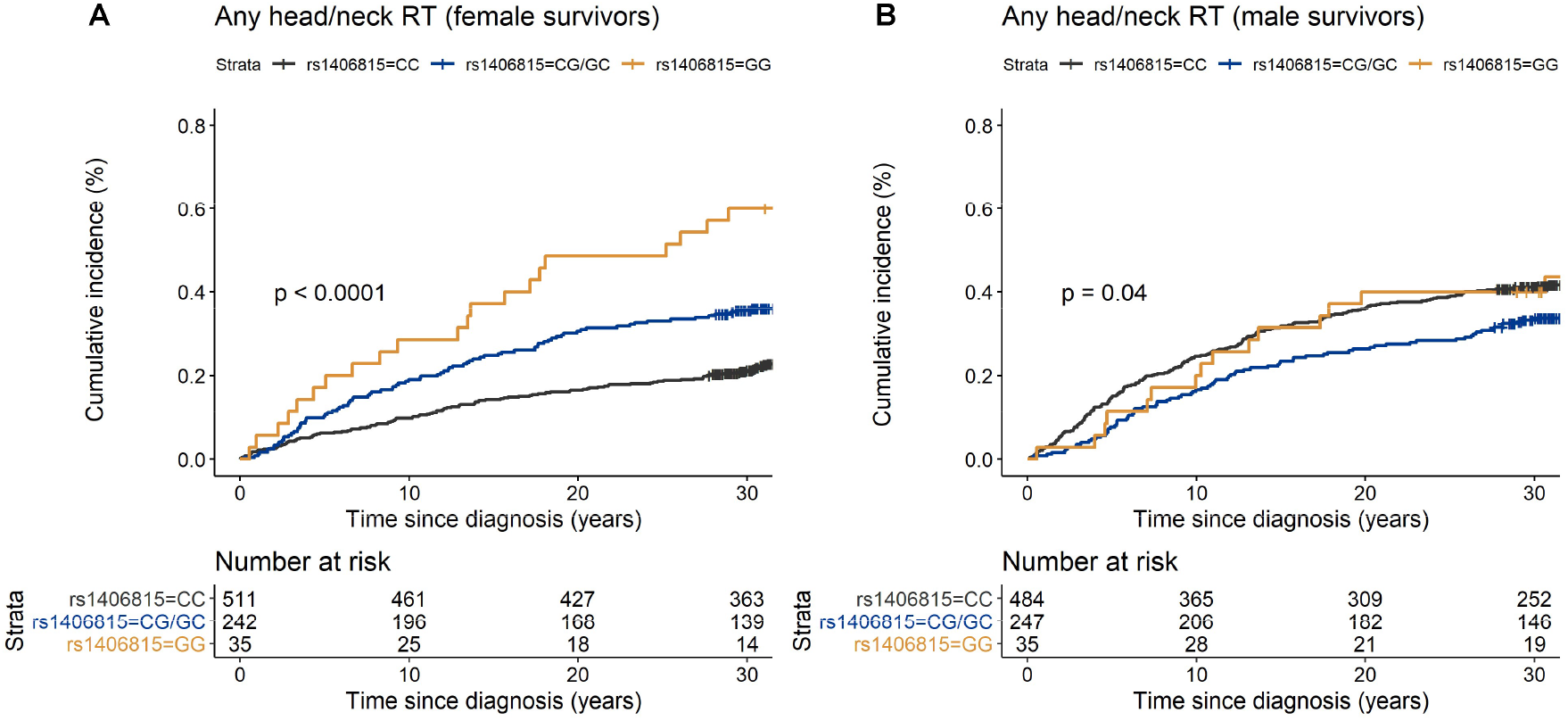
Cumulative incidence curves of post-diagnosis fracture in the CCSS female and male survivors with any exposure to head/neck radiation therapy (RT) by *HAGHL* locus SNP rs1406815 genotype profiles. Panel A shows cumulative incidence curves for fracture by SNP genotype among female survivors with any head/neck RT (N=788) while panel B is the corresponding figure among males (N=766). The fracture risk allele for SNP rs1406815 is allele G. The p-value from the two-sided log-rank test comparing the fracture risk probability distributions by genotype is provided in the lower left corner.

### *HAGHL* locus SNPs have plausible functional and regulatory consequences on fracture risk

We used external genomic annotation resources to interrogate *HAGHL* locus SNPs in a 99% credible set representing the set of common variants most likely to be responsible for the fracture risk association signal at the *HAGHL* locus. The 99% credible set for *HAGHL* locus SNPs consisted of 11 variants spanning a ∼14-kb region (Supplemental Table 6).

A PheWAS of UK Biobank phenotypes showed the top (P<1×10^−16^) associated phenotypes for credible-set SNPs were for height and body composition (Supplemental Table 8). While not phenome-wide significant, the leading (P<5×10^−3^) phenotypes for credible-set SNPs in a second PheWAS of ICD9 codes were largely related to musculoskeletal conditions. We also examined published GWAS of bone phenotypes conducted in the general population^(19,20)^, and found multiple credible-set SNPs were associated with nominally significant (P<0.05) decreases in estimated BMD and all showed non-significant but directionally consistent increases with fracture risk (Supplemental Table 9).

To determine likely gene targets and cellular contexts, we considered external functional annotations of the credible-set SNPs. Six SNPs mapped to *HAGHL* transcripts, of which two SNPs were also putative *HAGHL* coding alleles^(42)^ (rs1406815, encoding p.Arg50Gly; rs12448432, encoding p.[Ala202Thr;Ala94Thr;Ala84Thr;Ala21Thr]) (Supplemental Table 6). All credible-set SNPs’ fracture risk alleles were strongly associated with increased expression of *NARFL* and *HAGHL* (FDR≤5%), particularly in thyroid cells^(43)^ (Supplemental Table 6). Risk alleles for multiple credible-set SNPs (7/11) were also significantly associated (FDR<5%) with increased DNA methylation at a CpG site near *NARFL* and *HAGHL* (cg27144592) in whole blood.^(44)^ Lastly, among differential expression quantitative trait loci (eQTLs) in human osteoblasts treated with pharmacological agents known to affect bone cells^(45)^, rs12448432 was identified as a *cis*-eQTL (FDR<5%) for *HAGHL* in osteoblasts treated with dexamethasone or prostaglandin E2 compared to untreated samples.

We examined chromatin state annotations^(40)^ and found that credible-set SNPs predominantly overlapped putative promoter and transcribed regions (Supplemental Table 6). We therefore assessed whether the credible-set SNPs were likely to drive fracture risk signals in survivors by regulating promoter activity in specific cell types. We examined a set of four cell types likely to be relevant to fracture risk in female survivors exposed to head/neck RT and a comparison set of diverse cell types. We found that credible-set SNPs with high posterior probabilities (>0.2) for being causal fracture-risk SNPs co-localize with the *HAGHL* promoter region and showed significant selectivity (Bonferroni-corrected threshold P<3.6⨯10^−3^) for putative poised/bivalent promoter chromatin states in bone cells (osteoblasts, chondrocytes), female fetal brain tissue, and ovary tissue (Figure 4). No comparison cell type showed statistically significant enrichment of credible-set SNPs in promoter sites.

**Figure 4:**
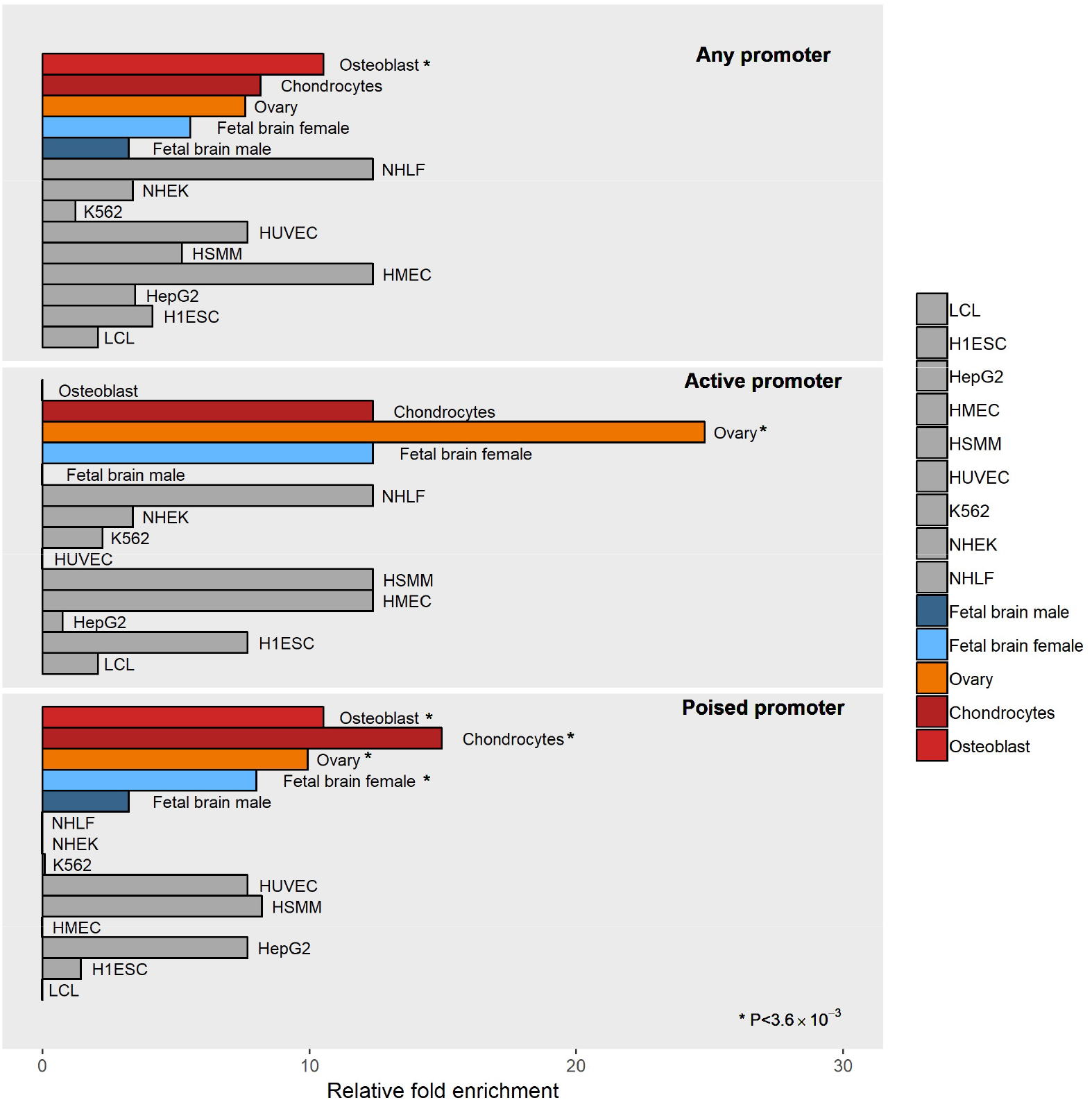
*HAGH*L/*NARFL* post-diagnosis fracture risk variants in female survivors overlap promoter epigenetic features in bone, ovary, and female brain cell types. Enrichments of posterior probabilities for credible-set SNPs that overlap promoter chromatin state annotations (25-state ChromHMM) compared to null distribution posterior probabilities are illustrated. Relative fold enrichments in 9 comparison ENCODE cell types are shown (gray), along with enrichments in 4 phenotype-relevant cell types, bone osteoblasts and chondrocytes (red), ovary (orange), and fetal brain cells from females (light blue), along with fetal brain cells from males (dark blue). Top, middle, and bottom panels show enrichment results from overlap with any promoter (active/poised promoters), active promoter, and poised promoter states, respectively. All significant enrichments are marked with * (Bonferroni-corrected threshold P<0.05/[14 evaluated cell types]=3.6×10^−3^). Abbreviations for ENCODE cell types are as follows: GM12878 (B-lymphocyte), K562 (chronic myelogenous leukemia), HepG2 (hepatocellular carcinoma), HSMM (skeletal muscle myoblast), HUVEC (umbilical vein endothelial), NHEK (epidermal keratinocyte), NHLF (lung fibroblast), H1-hESC (embryonic stem cell), HMEC (mammary epithelial]).

## DISCUSSION

In our GWAS of incident fracture risk after diagnosis in long-term survivors of childhood cancer, we identified seven novel genome-wide significant SNP associations at two genetic loci, *CD86* (3q13.33) and *HAGHL* (16p13.3), among 1,289 female survivors in CCSS. We confirmed three of the four fracture risk susceptibility SNPs at the *HAGHL* locus by replication in an independent cohort of 646 female survivors in SJLIFE. The fracture risk effects of replicated *HAGHL* locus SNPs increased incrementally in subsamples of female survivors with greater doses of RT to the head or neck, which was not observed among male survivors with comparable genotype and treatment profiles. In general population study samples, *HAGHL* locus SNPs had weak, non-significant associations with fracture risk,^(19)^ but nominal associations with BMD and genome-wide significant associations with skeletal size phenotypes (e.g., body height and mass)^(20,39)^. These results confirm that *HAGHL* credible set SNPs are plausible candidates for driving the observed association signal in female survivors, and further suggest that associated fracture risk effects may also depend on sex and RT exposures.

While evidence for increased risk for BMD deficits and fractures in survivors have been shown in literature, some studies have not observed greater-than-expected frequencies of bone-related late effects in survivors.^(1,18)^ Accordingly, current long-term follow-up guidelines for bone density and fracture late effects such as those issued by the Children’s Oncology Group^(46)^ are broad, recommending bone densitometry screening or clinical follow-up of all survivors with any exposure to radiation, antimetabolites, corticosteroids, or hematopoietic cell transplant. Our results suggest that an evaluation of both genetic and clinical risk factors and their interactions, i.e., *HAGHL* genetic variants modified by sex and varying exposures to head/neck RT, may potentially be more informative in identifying subgroups of female childhood cancer survivors at greater risk for fractures after diagnosis than existing follow-up recommendations.

Inconsistencies in the bone-related late effects literature also suggest that an improved understanding of the biological mechanisms underpinning fracture risk in survivors is warranted. Insight into how *HAGHL* locus SNPs affect fracture risk in female survivors, particularly those exposed to head/neck RT, may reveal new pathways for bone biology which are potentially useful as future targets for treatments for low bone density and osteoporosis. *In silico* analyses suggest that the fracture risk alleles at the *HAGHL* locus are strongly associated with *HAGHL* gene expression in endocrine tissues and may also play a role in the regulation of a poised/bivalent state at the *HAGHL* promoter in bone cell types. Notably, these *HAGHL* locus SNPs have also been associated with differential *HAGHL* gene expression in osteoblasts treated with dexamethasone and prostaglandin E2 (PGE2). *HAGHL* encodes a member of the glyoxalase II subfamily of the metallo-β-lactamase protein superfamily; while the exact functions of *HAGHL*-encoded glyoxalases remain unknown, glyoxalase I and II work in tandem in detoxifying pathways for byproducts of glycolysis and may contribute to the maturation of osteoclasts (bone-resorbing cells).^(47)^ Given that poised/bivalent promoter regions are posited as keeping genes “poised” for rapid activation in response to environmental stimuli, we speculate that *HAGHL* locus SNPs may increase fracture risk in female survivors by affecting osteoclastogenesis pathways mediated by *HAGHL* gene expression in response to head/neck radiation. PGE2 levels increase as a part of the local proinflammatory response after irradiation^(48)^ and increase in osteoblasts as an indirect effect of altered levels of thyroid hormones.^(49)^ Since damage to the thyroid gland and consequent thyroid hormone imbalance are common after head/neck RT^(35)^, head/neck RT may elevate fracture risk in female survivors with *HAGHL* risk alleles by altering baseline levels of thyroid hormones and PGE2 to influence *HAGHL* transcription and increase bone resorption.

To our knowledge, this study constitutes the first genome-wide assessment of SNP associations with fracture risk in long-term survivors of childhood cancer. Among the major strengths of our analysis is that we focus on capturing genetic variants with sex- and treatment-specific fracture risk effects in survivors. Furthermore, our analysis was restricted to incident fractures after primary cancer diagnosis instead of studying any lifetime fracture risk, the typical case definition for fracture in GWAS conducted in the general population. We also considered most major clinical risk factors used in fracture risk calculators^(50)^ (e.g., sex, age, height, weight) in our statistical models, as well as an extensive set of cancer treatments. However, our study has several limitations. Fractures were self-reported and are therefore subject to recall bias. While it would be ideal to study fractures confirmed by radiographic reports, previous studies of fracture risk in survivors suggest the validity of self-reported fractures is high.^(18)^ Because detailed fracture histories were only available for survivors who responded to a single CCSS follow-up questionnaire, these results may not be generalizable to all 5-year survivors. Secondary evaluations of low-trauma fractures could not be performed since data to distinguish traumatic fractures was unavailable. Temporal data for potential confounders at fracture occurrence were also unavailable, including for use of medications and supplements to improve BMD (e.g., hormone replacement therapy, vitamin D, calcium), alcohol use, smoking, exercise, height, and weight in CCSS; consequently, we did not account for these risk factors or used best available proxies (i.e., attained height/weight). While these risk factors are important contributors to fracture risk and would be useful in models for predicting fracture risk, there is evidence that adjusting for some of these factors may have limited impact on the results of GWAS of fracture risk.^(21)^ Lastly, posited biological mechanisms involving *HAGHL* locus SNPs, head/neck RT, and fracture risk in female survivors require functional validation, including an assessment of how these SNPs affect BMD.

In summary, we performed GWAS of first fracture risk following primary cancer diagnosis in long-term survivors of childhood cancer. We identified a credible novel genetic locus (*HAGHL*, 16p13.3) for fracture risk that is both female-specific and sensitive to previous exposures to head/neck RT. Our study demonstrates the importance of interrogating sex-specific SNP effects in survivors, especially for bone phenotypes with differential risk by sex in both the general population, due to sex-specific patterns for bone accretion and loss,^(7,8)^ and survivors, as a consequence of sex-specific vulnerabilities to cancer treatments.^(11,17)^ Because multiple clinical interventions to lessen fracture risk and increase BMD exist, future investigations to evaluate whether top genetic associations identified among survivors, including *HAGHL* genetic variants, can improve the prediction of future fracture risk in survivors should be pursued.

## Data Availability

The CCSS data used in this study may be accessed from the database of Genotypes and Phenotypes (dbGaP; https://www.ncbi.nlm.nih.gov/gap/) under accession number phs001327.v1.p1. Data from SJLIFE is available from the St. Jude Cloud (https://www.stjude.cloud/) under accession number SJC-DS-1002.

## ACKNOWLEDGEMENTS

This work was supported by the National Cancer Institute (U24 CA55727 to G.T.A., principal investigator, U01 CA195547 to M.M.H. and L.L.R., principal investigators, CA21765 to C. Roberts, Principal Investigator and R01 CA216354 to Y.Y. and J. Zhang principal investigators); Childhood Cancer Survivor Study Career Development Award; and American Lebanese Syrian Associated Charities.

Authors’ roles: C.I., Y.Y., S.B., G.T.A. designed study concept and analytic methodologies. C.L.W., K.K.N., W.C., S.B. informed clinical models. C.I., W.M. performed analyses. W.M.L., R.M.H., L.M.M., G.T.A., Z.W., W.C., K.K.N., C.L.W., Y.S., W.M., M.M.H., L.L.R. oversaw recruitment, sample collection, genotyping/sequencing, and data processing in CCSS and SJLIFE studies. C.I., N.L., Q.L., K.K.N. managed phenotype, clinical data. C.I., Y.Y. drafted the paper. All authors critically revised and approved the final manuscript. C.I. takes responsibility for the integrity of the data analysis.

